# Immediate Effects of COVID-19 Outbreak on Psychiatric Outpatients: Posttraumatic Stress and Influencing Factors

**DOI:** 10.1101/2020.08.12.20173468

**Authors:** Burç Çağrı Poyraz, Cana Aksoy Poyraz, Şenol Turan, Ömer Faruk Demirel, Yasin Kavla, Ersel Bulu, İrem Hacısalihoğlu, Elif Burcu Ersungur

## Abstract

We aimed to investigate the effects of COVID-19 outbreak and public health measures on the psychological well-being of patients with psychiatric disorders. This cross-sectional study assessed 436 outpatients recruited from a tertiary psychiatry clinic in Istanbul, Turkey, nearly one month after the government introduced strict measures of lockdown against the ongoing outbreak. Respondents completed a web-based survey on sociodemographic data, subjective sleep quality, and a range of psychiatric symptoms using the Impact of Events Scale-Revised (IES-R), and Hospital Anxiety and Depression Scale (HADS). Respondents reported high frequencies of clinically significant posttraumatic stress disorder (PTSD) (32.6%, IES-R score ≥ 33), anxiety (36.4%, HADS anxiety score > 10), and depression (51%, HADS depression score > 10). 20.5% of respondents described that their psychological status worsened during the COVID-19 outbreak, and 12.1% of respondents described poor or very poor sleep in the prior month. Positive predictors of increased PTSD symptoms included the chronic medical diseases, knowing someone in the social vicinity diagnosed with the COVID-19 infection, job loss or being on temporary leave after the outbreak, and increased exposure time to TV or social media. In contrast, male gender, older age, higher educational attainment, and the psychiatric diagnoses of schizophrenia and (to a lesser degree) bipolar disorder were the negative predictors. Our results suggest that patients with psychiatric disorders are prone to substantial psychological distress during the COVID-19 outbreak, and various individual, behavioral, and social factors mediate this effect.

## 1. Introduction

The 2019 novel coronavirus disease (COVID-19) emerged in China in December 2019 and spread worldwide rapidly. In Turkey, the first confirmed case was identified on March 10, 2020, and since then, plenty of regulations from the government have come into effect. Starting as of March 16, 2020, schools, universities, and public places, such as restaurants, cafes, and gyms were closed. On March 21, 2020, the Ministry of Interior announced a total curfew for those who were over 65 years of age or had chronic illnesses. On April 3, 2020, the curfew was further extended to people younger than 20 years. On April 10, 2020, a total lockdown during the weekends was placed in the provinces with metropolitan status. As of June 18, 2020, Turkey was one of the heavily hit countries by the COVID-19 pandemic, with 184.031 infected cases and 4882 people deceased. Istanbul, home to 16 million people has become the ‘epicenter’ of the outbreak in Turkey, and around 60% of the nationwide infections were documented in this city (Republic of Turkey Ministry of Health, 2020; “COVID-19 pandemic in Turkey,” n.d.)

As the COVID-19 pandemic rapidly sweeps around the world, it is inducing a massive impact on the institutions, economies, and healthcare systems, as well as the individuals at large. Recent studies have investigated the mental-health effects of the COVID-19 pandemic on society, with detrimental psychological consequences noted in a broad range of populations (Vindegaard and Benros, 2020). Yet, the mental health effects of the COVID-19 pandemic on the vulnerable groups, including the patients with psychiatric disorders, remain largely unaddressed. Identification of the mental health consequences of the COVID-19 pandemic in vulnerable populations might help to implement the timely and accurate psychosocial interventions to combat the negative effects in future.

In this study, we aimed to assess the symptoms of posttraumatic stress disorder (PTSD), anxiety and depression among the outpatients of a tertiary psychiatry clinic in the city of Istanbul, at the height of the COVID-19 outbreak. We also investigated the potential factors predicting the severity of PTSD symptoms. Another area of investigation was the difficulties met by the patients in using psychiatric services during the outbreak.

## 2. Methods

We conducted an online cross-sectional survey among the outpatients of the department of psychiatry at Cerrahpaşa Medical Faculty in Istanbul, Turkey. The online survey was designed using the Survey Monkey website, and invitations to participate in the study were sent via WhatsApp messages. The study was conducted between 19 April and 19 May 2020, when the lockdown regulations were still in effect, and the daily new cases across the country varied between 1022 and 4674.

An electronic informed consent form was presented on the first page of the survey citing the purposes and the voluntary nature of the survey, and that all information provided by the participants would be kept confidential, and they could withdraw from the survey at any time. The procedures of this study complied with the provisions of the Declaration of Helsinki concerning research on Human participants. The Ethics Committee of Cerrahpaşa Medical Faculty approved this study.

### 2.1. Subjects

We sent online invitations to each patient who had visited the above department’s outpatient clinics in the preceding three months and who were aged 18 years or above and who could be contacted via WhatsApp messages. Target populations were the outpatients of the general psychiatry, consultation-liaison and geriatric psychiatry divisions, and bipolar and psychotic disorders units.

### 2.2. The Online Survey

The online survey contained three parts and 63 questions. The first part included the questionnaire on sociodemographic data (age, gender, educational, marital and employment status, and household size) and additional information (having a child below 18 years of age, the presence of an individual above 65 years of age in the household, chronic medical diseases, personal/family history of COVID-19 infection, history of COVID-19 infection in respondent’s relatives, friends and acquaintance, change in respondent’s employment status during the outbreak, respondent’s main source of information on the COVID-19 pandemic, duration of daily TV and social media exposure, questions on whether the respondent was experiencing difficulty in reaching the psychiatric services during the outbreak, and respondent’s view on seriousness of the COVID-19 pandemic).

The second and third parts of the survey included the evaluation of psychological distress. Symptoms of PTSD related to the outbreak were assessed using the Impact of Event Scale-Revised (IES-R) (Weiss and Marmar, 1997). This tool is a 22-item self-report questionnaire that evaluates symptoms of intrusion, avoidance and hyperarousal, and presents a total score for the subjective stress related to a traumatic event. IES-R was frequently used after a variety of traumatic settings (Morina et al., 2013), and after major public health crises (Lee et al., 2018; Varshney et al., 2020; Wang et al., 2020). We made slight modifications to the Turkish adaptation of the IES-R (Corapcioglu et al., 2006) (replacing the word ‘event’ with ‘outbreak’, where appropriate) in order to account for the nature of the event investigated. Anxious and depressive symptoms were assessed using the Hospital Anxiety and Depression Scale (HADS) (Zigmond and Snaith, 1983), a 14-item self-report questionnaire. HADS can assess symptom severity and caseness of anxiety and depressive disorders, and it consists of anxiety and depression subscales. HADS was validated in a variety of populations, including the general medical and community settings (Bjelland et al., 2002; Bocéréan and Dupret, 2014; Djukanovic et al., 2017). Both IES-R and HADS were validated previously for the Turkish population (Corapcioglu et al., 2006; Aydemir et al., 1997). Respondent’s rating of his/her sleep quality in the prior month and of the change in his/her psychological status during the outbreak was assessed using Likert-type questions. The estimated time to complete the survey was from 15 to 20 min.

### 2.3. Statistical Analysis

Descriptive statistics were calculated for sociodemographic characteristics and additional data. The scores of the IES-R and HADS subscales were expressed as mean and standard deviation. We used linear regressions to calculate the univariate associations between sociodemographic characteristics and additional data, and IES-R, HADS anxiety, and HADS depression scores. We also performed multiple linear regression analysis for predicting the severity of PTSD symptoms using the IES-R scores as the outcome variable, and selected significant variables as independent predictors. We adjusted this analysis for age, gender, and education status. All tests were two-tailed, with a significance level of p < 0.05. Statistical analysis was performed using SPSS Statistic 23.0 (IBM SPSS Statistics, New York, United States).

## 3. Results

### 3.1. Survey Respondents

We sent a total of 2700 survey messages to the outpatients via WhatsApp, and 485 responses were received (response rate = 18%). On average, respondents spent 16 minutes on the survey. Four hundred thirty-six surveys with a complete IES-R were included in the analysis. IES-R, HADS anxiety, and HADS depression exhibited high internal consistency in our sample: Cronbach’s alphas for these scales were 0.917, 0.895, and 0.805, respectively.

The psychological impact of the COVID-19 outbreak on our sample as measured by the IES-R revealed a mean sample score of 26.7 (SD=15.3; range=0-74). In our sample, 142 respondents (32.6%) met the ‘probable’ diagnosis of PTSD (IES-R score equal to or above 33 (Creamer et al., 2003)). Mean sample scores for HADS anxiety and HADS depression were 8.8 (SD=5.6; range=0-21) and 10.6 (SD=4.7; range=0-21), respectively. Identification of’probable’ cases of anxiety and depression was based on the cutoff scores recommended by the authors of the HADS scale (Zigmond and Snaith, 1983). One hundred fifty-seven respondents (36.4%) were considered having ‘probable’ anxiety (HADS anxiety score above 10), and 220 respondents (51%) were considered having ‘probable’ depression (HADS depression score above 10). When we used a lower cutoff score for the depression subscale as recommended by the Turkish adaptation study (HADS depression score above 7 (Aydemir et al., 1997)), 318 respondents (73.8%) had ‘probable’ depression.

### 3.2. Sociodemographic Characteristics and Psychological Impact (Table 1)

The majority of respondents were female (67.4%), between 18-29 years of age (33.1%), married (42.2%), employed (48.1%), and had a university or higher education (42.1%). Overall, the male gender was significantly associated with lower scores in the IES-R (β=-0.17, p<0.01) and HADS anxiety (β=-0.13, p<0.01). Respondents’ age at 40-49 or below was significantly associated with higher scores in HADS anxiety and HADS depression as compared to respondents’ age of 60 and above. Lower-secondary school education was significantly associated with higher IES-R (β=0.10, p<0.05) and HADS depression scores (β=0.12, p<0.05).

**Table 1.**
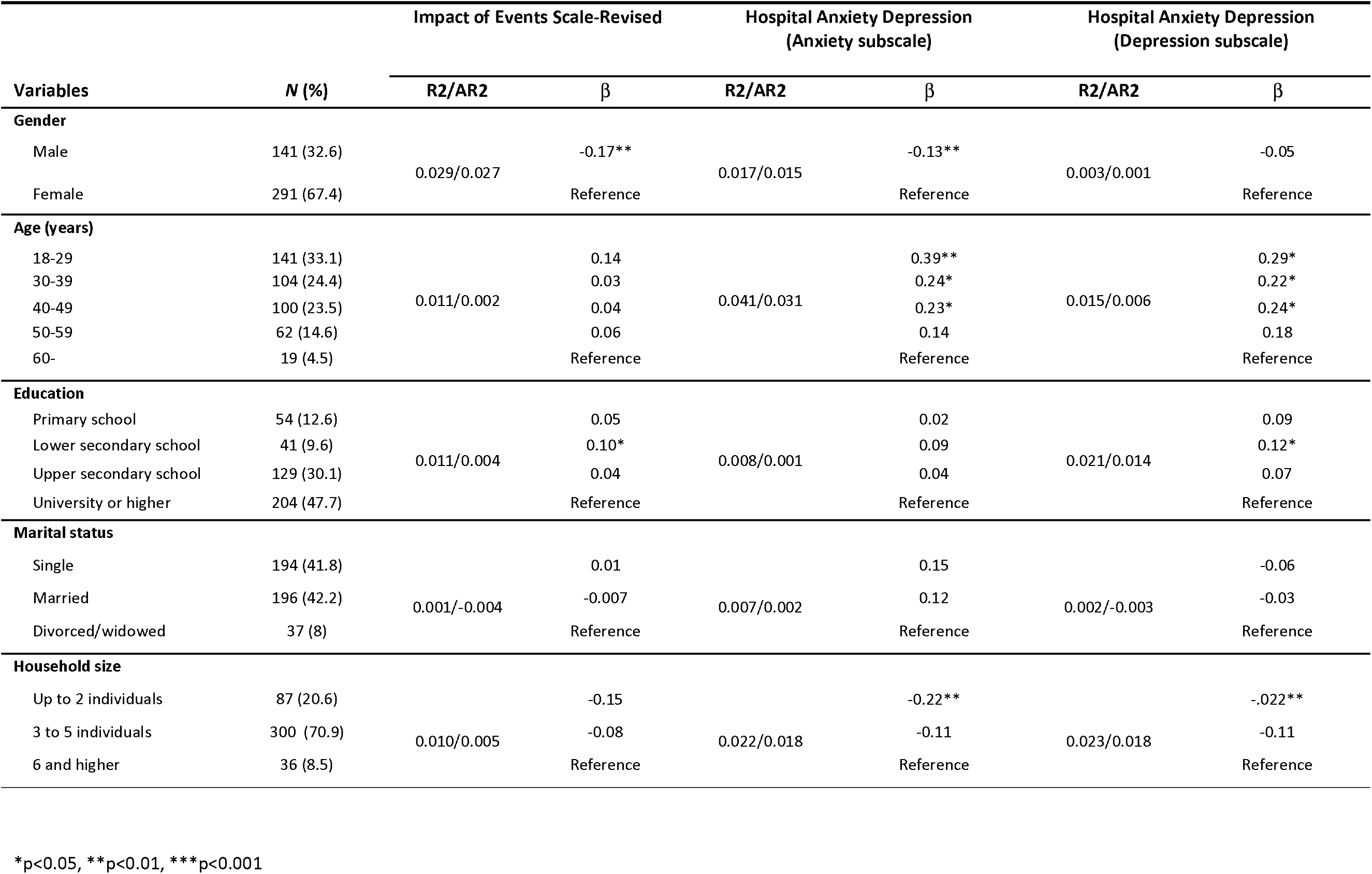
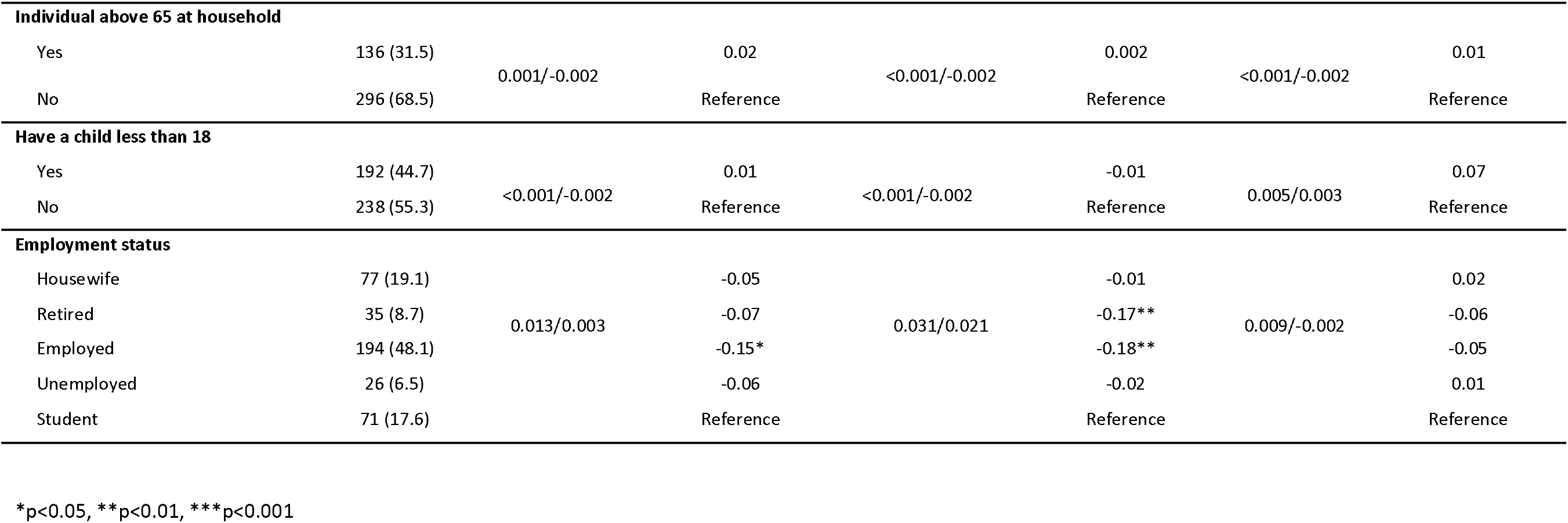
Sociodemographic chareacteristics with symptoms of posttraumatic stress, anxiety and depression.

|  |  | Impact of Events Scale-Revised |  | Hospital Anxiety Depression<br>(Anxiety subscale) |  | Hospital Anxiety Depression<br>(Depression subscale) |  |
| --- | --- | --- | --- | --- | --- | --- | --- |
| Variables | N (%) | R2/AR2 | β | R2/AR2 | β | R2/AR2 | β |
| Gender |  |  |  |  |  |  |  |
| Male | 141 (32.6) | 0.029/0.027 | -0.17** | 0.017/0.015 | -0.13** | 0.003/0.001 | -0.05 |
| Female | 291 (67.4) |  | Reference |  | Reference |  | Reference |
| Age (years) |  |  |  |  |  |  |  |
| 18-29 | 141 (33.1) | 0.011/0.002 | 0.14 | 0.041/0.031 | 0.39** | 0.015/0.006 | 0.29* |
| 30-39 | 104 (24.4) |  | 0.03 |  | 0.24* |  | 0.22* |
| 40-49 | 100 (23.5) |  | 0.04 |  | 0.23* |  | 0.24* |
| 50-59 | 62 (14.6) |  | 0.06 |  | 0.14 |  | 0.18 |
| 60- | 19 (4.5) |  | Reference |  | Reference |  | Reference |
| Education |  |  |  |  |  |  |  |
| Primary school | 54 (12.6) | 0.011/0.004 | 0.05 | 0.008/0.001 | 0.02 | 0.021/0.014 | 0.09 |
| Lower secondary school | 41 (9.6) |  | 0.10* |  | 0.09 |  | 0.12* |
| Upper secondary school | 129 (30.1) |  | 0.04 |  | 0.04 |  | 0.07 |
| University or higher | 204 (47.7) |  | Reference |  | Reference |  | Reference |
| Marital status |  |  |  |  |  |  |  |
| Single | 194 (41.8) | 0.001/-0.004 | 0.01 | 0.007/0.002 | 0.15 | 0.002/-0.003 | -0.06 |
| Married | 196 (42.2) |  | -0.007 |  | 0.12 |  | -0.03 |
| Divorced/widowed | 37 (8) |  | Reference |  | Reference |  | Reference |
| Household size |  |  |  |  |  |  |  |
| Up to 2 individuals | 87 (20.6) | 0.010/0.005 | -0.15 | 0.022/0.018 | -0.22** | 0.023/0.018 | -.022** |
| 3 to 5 individuals | 300 (70.9) |  | -0.08 |  | -0.11 |  | -0.11 |
| 6 and higher | 36 (8.5) |  | Reference |  | Reference |  | Reference |

\*p<0.05, \*\*p<0.01, \*\*\*p<0.001

Table 1 (Continued). Sociodemographic characteristics, and their associations with symptoms of posttraumatic stress, anxiety and depression.
|  |  | Impact of Events Scale-Revised |  | Hospital Anxiety Depression<br>(Anxiety subscale) |  | Hospital Anxiety Depression<br>(Depression subscale) |  |
| --- | --- | --- | --- | --- | --- | --- | --- |
| Variables | N (%) | R2/AR2 | β | R2/AR2 | β | R2/AR2 | β |
| Individual above 65 at household |  |  |  |  |  |  |  |
| Yes | 136 (31.5) | 0.001/-0.002 | 0.02 | <0.001/-0.002 | 0.002 | <0.001/-0.002 | 0.01 |
| No | 296 (68.5) |  | Reference |  | Reference |  | Reference |
| Have a child less than 18 |  |  |  |  |  |  |  |
| Yes | 192 (44.7) | <0.001/-0.002 | 0.01 | <0.001/-0.002 | -0.01 | 0.005/0.003 | 0.07 |
| No | 238 (55.3) |  | Reference |  | Reference |  | Reference |
| Employment status |  |  |  |  |  |  |  |
| Housewife | 77 (19.1) | 0.013/0.003 | -0.05 | 0.031/0.021 | -0.01 | 0.009/-0.002 | 0.02 |
| Retired | 35 (8.7) |  | -0.07 |  | -0.17** |  | -0.06 |
| Employed | 194 (48.1) |  | -0.15 * |  | -0.18** |  | -0.05 |
| Unemployed | 26 (6.5) |  | -0.06 |  | -0.02 |  | 0.01 |
| Student | 71 (17.6) |  | Reference |  | Reference |  | Reference |

\*p<0.05, \*\*p<0.01, \*\*\*p<0.001

Employed status was significantly associated with lower IES-R (β=-0.15, p<0.05) and HADS anxiety scores (β=-0.18, p<0.01) compared to student status. Also, retired status was significantly associated with lower HADS anxiety scores (β=-0.17, p<0.01). Household size up to 2 individuals was significantly associated with lower HADS anxiety and HADS depression scores (β=-0.22, p<0.01; each, respectively). Other sociodemographic characteristics, including marital status, having a child under 18 and the presence of an individual above 65 in the household were not associated with the IES-R and HADS subscales.

### 3.3. Psychiatric Diagnosis, Chronic Medical Disease, Sleep quality and Psychological Impact (Table 2)

In our sample, 110 respondents (25.2%) had unipolar depression, 81 respondents (18.6%) had an anxiety spectrum disorder (i.e., panic disorder, generalized anxiety disorder, mixed anxiety and depressive disorder or obsessive-compulsive disorder), 45 respondents (10.3%) had the diagnosis of schizophrenia, 44 respondents (10.1%) had bipolar disorder, and 33 respondents (7.6%) had a psychiatric diagnosis other than the above categories (i.e., somatoform disorders, eating disorders, adult-type ADHD, severe personality disorders or multiple diagnoses), as confirmed by review of the outpatient files. 123 respondents (28.2%) did not disclose their identities in the survey, and we analyzed these subjects as a separate group (‘unspecified’).

**Table 2.** Psychiatric Diagnosis, Medical Comorbidity, Sleep quality and their associations with symptoms of posttraumatic stress, anxiety and depression.

|  |  | Impact of Events Scale-Revised |  | Hospital Anxiety Depression<br>(Anxiety subscale) |  | Hospital Anxiety Depression<br>(Depression subscale) |  |
| --- | --- | --- | --- | --- | --- | --- | --- |
| Variables | N (%) | R2/AR2 | β | R2/AR2 | β | R2/AR2 | β |
| Chronic medical disease |  |  |  |  |  |  |  |
| One or more disease(s) | 127 (68.9) | 0.015/0.012 | 0.12* | 0.005/0.003 | 0.07 | 0.010/0.008 | 0.10* |
| None | 281 (31.1) |  | Reference |  | Reference |  | Reference |
| Psychiatric diagnosis |  |  |  |  |  |  |  |
| Unipolar depression | 110 (25.2) | 0.110/0.100 | -0.004 | 0.091/0.080 | -0.001 | 0.058/0.047 | -0.005 |
| Bipolar disorder | 44 (10.1) |  | -0.15** |  | -0.17** |  | -0.13* |
| Schizophrenia | 45 (10.3) |  | -0.32*** |  | -0.27*** |  | -0.22*** |
| Other | 33 (7.6) |  | -0.02 |  | -0.01 |  | 0.02 |
| Unspecified | 123 (28.2) |  | -0.15* |  | -0.14* |  | -0.07 |
| Anxiety disorder | 81 (18.6) |  | Reference |  | Reference |  | Reference |
| How did your psychological status change during COVID outbreak? |  |  |  |  |  |  |  |
| “Not know” | 37 (8.6) | 0.272/0.266 | 0.12** | 0.236/0.230 | 0.03 | 0.223/0.217 | 0.13** |
| “Very much improved” / “much improved” | 32 (7.4) |  | -0.14** |  | -0.17*** |  | -0.17*** |
| “Minimally improved” / “No change” / “minimally worse” | 273 (63.4) |  | Reference |  | Reference |  | Reference |
| “Much worse” / “very much worse” | 88 (20.5) |  | 0.48*** |  | 0.43*** |  | 0.40*** |
| How was your sleep quality during last month? |  |  |  |  |  |  |  |
| Very good | 56 (13) | 0.262/0.252 | -0.31*** | 0.245/0.234 | -0.33*** | 0.226/0.215 | -0.34*** |
| Good | 99(23) |  | -0.33*** |  | -0.31*** |  | -0.24*** |
| Fairly good | 79 (18.3) |  | -0.15** |  | -0.11* |  | -0.10* |
| Average | 109 (25.3) |  | Reference |  | Reference |  | Reference |
| Fairly poor | 36 (8.4) |  | 0.07 |  | 0.06 |  | 0.08 |
| Poor | 28 (6.5) |  | 0.12** |  | 0.11* |  | 0.08 |
| Very poor | 24 (5.6) |  | 0.17*** |  | 0.15** |  | 0.18*** |

\* $p < 0.05$ , \*\* $p < 0.01$ , \*\*\* $p < 0.001$ . <sup>a</sup> Overall, 38 respondents had hypertension, 42 respondents had diabetes mellitus, 27 respondents had cardiovascular disease, 24 respondents had chronic pulmonary disease, 18 respondents had been diagnosed with cancer, 35 respondents had a rheumatological disease, inflammatory bowel disease or multiple sclerosis, and 31 had an endocrine disorder.

In our sample, the diagnosis of schizophrenia and (to a lesser degree) the diagnosis of bipolar disorder were significantly associated with lower scores in the IES-R (β=-0.32, p<0.001 and β=-0.15, p<0.01, respectively), HADS anxiety (β=-0.29, p<0.001 and β=-0.18, p<0.01, respectively), and HADS depression (β=-0.24, p<0.001 and β=-0.14, p <0.01, respectively). Overall, 44.4% of patients with anxiety spectrum disorders, 43.5% of patients with unipolar depression, 18.2% of patients with bipolar disorder, 2.2% of patients with schizophrenia, and 45.5% of patients with a diagnosis other than the above categories met the ‘probable’ diagnosis of PTSD (IES-R score equal to or above 33). 27.6% of patients in the ‘unspecified’ group had ‘probable’ PTSD.

Two hundred seventy-three respondents (63.4%) reported that they did not experience a major change in their psychological status during the COVID-19 outbreak, while 88 respondents (20.5%) reported that their psychological status worsened significantly (a self-rating of “worse” or “very much worse”). Conversely, 32 respondents (7.4%) reported that their psychological status improved “much” or “very much”. In our sample, respondents’ self-report of worsening psychological status was significantly associated with higher IES-R, HADS anxiety, and HADS depression scores (β=0.48, p<0.001; β=0.43, p<0.001 and β=0.40, p<0.001, respectively). In contrast, respondents’ report of improvement in psychological status was significantly associated with lower IES-R, HADS anxiety and HADS depression scores (β=-0.14, p<0.01; β=-0.17, p<0.001 and β=-0.17, p<0.001, respectively). A chi-square test (self-report of worsening psychological status X psychiatric diagnosis) showed that reported worsening of the psychological status differed by diagnostic groups (X^2^ (5, *N* = 430) = 25.02, p<0.001), and a significantly lower number of patients with schizophrenia reported worsening of their psychological status (p<0.05, post hoc analyses with Bonferroni correction).

Fifty-two respondents (12.1%) reported ‘poor’ or ‘very poor’ sleep in the prior month, and this was significantly associated with higher IES-R, HADS anxiety, and HADS depression scores. Additionally, having a chronic medical disease was significantly associated with higher scores in the IES-R (β=0.12, p<0.05) and HADS depression (β=0.10, p<0.05).

### 3.4. Public and Media Exposure to COVID-19, Employment Status Change and Psychological Impact (Table 3)

Five respondents (1.2%) reported that they had been diagnosed with the COVID-19 infection, while 11 respondents (2.55%) reported a history of COVID-19 infection in an immediate family member. Forty-six respondents (10.6%) had a relative, 26 respondents (6%) had a close friend, and 77 respondents (17.8%) had an acquaintance who had the infection in the recent past. Overall, 136 respondents (31.5%) had one or more individuals in their social vicinity who contracted the COVID-19 infection. Knowing someone in the social vicinity diagnosed with COVID-19 was significantly associated with higher scores in the IES-R (β=0.14, p<0.01), HADS anxiety (β=0.14, p<0.01), and HADS depression (β=0.09, p<0.05).

**Table 3.** Public and Media Exposure to COVID-19, Employment Status Change and their associations with symptoms of posttraumatic stress, anxiety and depression.

|  |  | Impact of Events Scale-Revised |  | Hospital Anxiety Depression<br>(Anxiety subscale) |  | Hospital Anxiety Depression<br>(Depression subscale) |  |
| --- | --- | --- | --- | --- | --- | --- | --- |
| Variables | N (%) | R2/AR2 | β | R2/AR2 | β | R2/AR2 | β |
| COVID infection in the social vicinity |  |  |  |  |  |  |  |
| Present | 136 (31.5) | 0.021/0.019 | 0.14** | <0.021/0.010 | 0.14** | <0.009/0.007 | 0.09* |
| Absent | 296 (68.5) |  | Reference |  | Reference |  | Reference |
| How serious do you view the COVID pandemic? |  |  |  |  |  |  |  |
| “not a real threat” | 14 (3.2) | 0.035/0.028 | 0.04 | 0.019/0.012 | 0.06 | 0.020/0.013 | 0.02 |
| “a small threat” | 44 (10.2) |  | Reference |  | Reference |  | Reference |
| “a serious threat” | 156 (36.1) |  | -0.01 |  | 0.06 |  | 0.02 |
| “a very serious threat” | 219 (50.7) |  | 0.17* |  | 0.18* |  | 0.16 |
| Employment status change during COVID outbreak |  |  |  |  |  |  |  |
| No change | 58 (14.1) | 0.022/0.013 | Reference | 0.006/-0.004 | Reference | 0.026/0.017 | Reference |
| Job lost or temporary leave | 34 (8.4) |  | 0.14* |  | 0.06 |  | 0.12* |
| Started working from home or infrequent office visits | 82 (20) |  | 0.08 |  | 0.01 |  | -0.01 |
| Paid leave | 14 (3.4) |  | -0.03 |  | -0.04 |  | -0.08 |
| Not a worker | 224 (54.4) |  | 0.13 |  | 0.04 |  | 0.05 |
| Main source(s) of information on COVID |  |  |  |  |  |  |  |
| TV | 232 (56) | 0.011/0.002 | Reference | 0.014/0.004 | Reference | 0.011/0.002 | Reference |
| Newspapers | 9 (2.2) |  | -0.05 |  | -0.03 |  | -0.03 |
| Social media/Internet | 120 (29) |  | 0.06 |  | 0.10* |  | 0.04 |
| Mixed | 43 (10.4) |  | 0.05 |  | 0.05 |  | -0.06 |
| Other | 10 (2.4) |  | -0.03 |  | 0.01 |  | -0.05 |
| Daily exposure time to TV and social media |  |  |  |  |  |  |  |
| None | 10 (2.3) | 0.063/0.052 | 0.002 | 0.030/0.018 | 0.04 | 0.039/0.028 | 0.04 |
| Less than 2 hours | 83 (19.2) |  | -0.31*** |  | -0.14 |  | -0.23** |
| 2-3 hours | 174 (40.2) |  | -0.35*** |  | -0.17* |  | -0.27** |
| 4-5 hours | 95 (21.9) |  | -0.18* |  | -0.03 |  | -0.17* |
| 6-7 hours | 33 (7.6) |  | -0.11 |  | -0.002 |  | -0.12* |
| 8 hours and above | 38 (8.8) |  | Reference |  | Reference |  | Reference |

\*p<0.05, \*\*p<0.01, \*\*\*p<0.001

TV (56%) and social media (29%) were reported as the main sources of information on the COVID-19 outbreak in our sample, and for 78.5% of the respondents, daily TV and/or social media exposure time was at 2-3 hours or above. Social media as the main source of information was significantly associated with higher scores in HADS anxiety, when compared to TV as the main source (β=0.10, p<0.05). In our sample, decreased daily exposure time to TV and social media was significantly associated with lower scores on the IES-R, HADS anxiety, and HADS depression.

Thirty-four respondents (8.4%) in our sample reported that they lost their job or were on temporary leave after the COVID-19 outbreak. Job loss or being on temporary leave after the COVID-19 outbreak was significantly associated with higher scores in the IES-R (β=0.14, p<0.05) and HADS depression (β=0.12, p<0.05).

Respondents’ perception of the COVID-19 outbreak as ‘a very serious’ threat was significantly associated with higher scores in the IES-R (β=0.17, p<0.05) and HADS anxiety (β=0.18, p<0.05).

### 3.5. Change in Respondents’ Use of Psychiatric Services during the COVID-19 Outbreak (Table 3)

One hundred twenty-nine (29.6%) respondents reported that they did not face a significant problem using psychiatric outpatient services, while 202 respondents (46.3%) reported that they avoided outpatient visits because of the fear of contracting the infection. Twenty-nine (6.6%) respondents reported that they were not able get an appointment at the outpatient clinic or their appointments were canceled, and 28 respondents (6.4%) reported that they were not able to get their prescriptions filled. According to 18 respondents (4.1%), their latest outpatient visit was short and unsatisfying. Overall, 57 respondents (13.7%) reported medication discontinuation during the outbreak. Linear regression analysis showed that avoiding the outpatient visits because of the fear of contracting the infection was significantly associated with higher scores in the IES-R (β=0.28, p<0.001), HADS anxiety (β=0.22, pcO.OOl), and HADS depression (β=0.23, p<0.001).

### 3.6. Prediction of COVID-19 Outbreak related PTSD Symptoms (Table 4)

Multiple linear regression model (F(12,349) = 8.63, p<0.001) with an R^2^ of 0.22 showed that chronic medical diseases, knowing someone in the social vicinity diagnosed with COVID-19 infection, job loss or being on temporary leave, and increased daily exposure time to TV and social media were the positive predictors of increased PTSD symptoms related to the outbreak. In contrast, the male gender, older age, higher educational attainment, and the psychiatric diagnoses of schizophrenia and bipolar disorder (compared to the other diagnostic categories) were the negative predictors.

**Table 4.** Multiple linear regression analysis of factors predicting posttraumatic stress.

| Model | Posttraumatic Stress (IES-R) |  |  |  |  |
| --- | --- | --- | --- | --- | --- |
|  | Unstandardized Coefficients |  | Standardized Coefficients |  |  |
| | B | Std. Error | Beta ($\beta$) | t | Sig. |
| **Male** | -4.19 | 1.48 | **-0.13** | -2.82 | **0.005** |
| **Age (decades)** | -2.21 | 0.63 | **-0.18** | -3.49 | **0.001** |
| **Education status** | -1.80 | 0.65 | **-0.13** | -2.75 | **0.006** |
| **Psychiatric diagnosis** |  |  |  |  |  |
| Unipolar depression | -0.52 | 2.04 | -0.01 | -0.25 | 0.79 |
| Bipolar disorder | -6.08 | 2.55 | **-0.13** | -2.38 | **0.01** |
| Schizophrenia | -11.98 | 2.61 | **-0.26** | -4.58 | **<0.001** |
| Other | 2.48 | 2.86 | 0.04 | 0.86 | 0.38 |
| Unspecified | -3.33 | 2.03 | -0.10 | -1.64 | 0.10 |
| Anxiety disorder | - | - | - | - | - |
| **Chronic Medical disease** | 4.96 | 1.59 | **0.16** | 3.11 | **0.002** |
| **COVID-19 infection in the social vicinity** | 3.75 | 1.47 | **0.12** | 2.55 | **0.01** |
| **Daily exposure time to TV or social media** | 1.30 | 0.56 | **0.11** | 2.30 | **0.02** |
| **Job lost or temporary leave during the outbreak** | 5.82 | 2.46 | **0.11** | 2.36 | **0.01** |

## 4. Discussion

We found that at the height of the COVID-19 outbreak in a metropolitan area, a high percentage of patients with psychiatric disorders experienced significant psychological distress. Specifically, around a third of the patients in our study met the ‘probable’ diagnosis of PTSD, with over 80% of these subjects reporting comorbid depression. Around one-third of the patients in our study had clinically significant anxiety, and half of the patients reported clinically significant depression. Also, over one-fifth of the patients felt that their psychological status worsened considerably during the COVID-19 outbreak, and over one-tenth reported poor sleep quality in the prior month.

Studies investigating the psychological effects of the epidemics and related public health measures on society have increased dramatically after the COVID-19 outbreak. Majority of recent studies examined the public at large, university students, health care workers, and quarantined and infected individuals, and these found a high level of psychological impact related to the outbreak: that is, elevated symptoms of posttraumatic stress, anxiety, and depression as well as impaired sleep across individuals (Brooks et al., 2020; Vindegaard and Benros, 2020).

Research on the psychiatric patients who are potentially among the most vulnerable to the adverse psychological effects of the pandemic, however, has been scarce. An online study that investigated 2065 psychiatric outpatients from the city of Chengdu, China found that around a quarter of the patients had clinically significant anxiety and insomnia, and one in six patients had depression (Zhou et al., 2020). The psychological distress reported was lower compared to our findings. This may be because the setting was a mildly affected area in China, and the study time coincided with the end of the epidemic in that country. This study also did not assess specific PTSD symptoms, which prevents ascribing the severity of existing psychiatric symptoms straightly to the outbreak. Yet, around 20% of the patients in the study reported worsening of their mental health because of the pandemic, which agrees with our findings. A similar online study from Italy, investigating 205 patients with serious mental illness found that at the peak of the outbreak, subjects had significantly higher levels of stress, anxiety, and depression compared to healthy controls (Iasevoli et al., 2020). This study also did not assess the specific PTSD symptoms. A third online study, however, compared the severity of symptoms of PTSD, anxiety, and depression between the psychiatric outpatients (N=76) and healthy controls, and identified a greater psychological impact of the outbreak on the former group (Hao et al., 2020). Notably, 31% of the patients in the study had a ‘probable’ diagnosis of PTSD (IES-R score equal to 24 or above), in contrast to only 13.8% of the controls. The frequency of the significant PTSD symptoms was higher in our sample: 54% of the patients could be identified as having ‘probable’ PTSD at this cutoff level.

We investigated the factors predicting increased PTSD symptoms in our sample. Of the sociodemographic variables, female gender, young age, and low educational attainment were the positive predictive factors of increased PTSD symptoms. In line with these results, earlier research reported that an important risk factor for PTSD was the female gender (Sareen, 2014). Similarly, adults with low educational attainment were reported to be at high risk for PTSD after the earthquakes (Tang et al., 2017). A recent review on the psychological impact of the COVID-19 pandemic concluded that depression and anxiety were more common in females and subjects with low educational attainment. Findings on the effects of age were inconsistent (Vindegaard and Benros, 2020).

We found that among the psychiatric conditions, unipolar depression and anxiety spectrum disorders were significantly related to high levels of PTSD symptoms. Conversely, the diagnoses of schizophrenia and, to a much smaller extent, bipolar disorder were related to the lesser severity of PTSD symptoms. A significantly lesser number of patients with schizophrenia reported worsening of their psychological status, and interestingly, only one patient with schizophrenia (2.2%) had the PTSD symptoms severe enough to indicate caseness. In contrast, earlier studies found that the occurrence of PTSD in schizophrenia was higher than the rate of PTSD in the general population, even after controlling for the amount of lifetime traumatic experiences (Lommen and Restifo, 2009; Dallel et al., 2018). There are a number of explanations that might account for the low frequency of PTSD symptoms in patients with schizophrenia in our study. One possibility is the relative underreporting of PTSD symptoms in patients with schizophrenia. Also, a lesser interruption in everyday activities of the patients with schizophrenia, compared to other diagnostic groups, might have decreased the impact of the outbreak and lockdown on these patients. A large number of patients with severe mental disorders live with their family members in Turkey (Özden and Tuncay, 2018), and this might have been also protective by increasing social support. Most interestingly, however, the resilience to the impact of the current pandemic may come from the severe negative symptoms in schizophrenia. This is supported by a recent report of an eight times lower risk of lifetime PTSD in the deficit syndrome of schizophrenia compared to the non-deficit syndrome. (Strauss et al., 2011). The authors of this study argued that diminished experience of negative emotions and cognitive impairments in schizophrenia may preclude these patients from re-experiencing salient traumatic events, and this may be protective against the PTSD in the deficit schizophrenia. Even though we were unable to determine the extent to which the patients with schizophrenia in our sample had the deficit syndrome, a high frequency is plausible. As for the bipolar disorder group, 18.2% of the subjects reported clinically significant PTSD symptoms. Although this frequency is lower than that was found in patients with anxiety spectrum disorders and unipolar depression, a substantial proportion of patients with bipolar disorder still met the ‘probable’ diagnosis of PTSD.

The presence of chronic medical diseases was independently related to the symptoms of PTSD (Table 4) and depression (data not shown). It can be argued that being aware of the significant mortality risk in relation to the COVID-19 infection in chronic medical diseases (that was widely reported by the media) might have put some subjects in a vulnerable position by increasing their infection-related fears. Alternatively, chronic medical diseases might predispose an individual to psychological distress by complex psychosocial and biological effects. In parallel, recent studies on the mental health effects of the COVID-19 pandemic reported an increased risk of depression and anxiety in subjects with chronic medical diseases (Vindegaard and Benros, 2020).

In this study, the number of subjects who reported knowing one or more individuals in their social vicinity (i.e., family members, relatives, friends, or acquaintance) contracting the COVID-19 infection was high (31.5%), and this was significantly associated with increased levels of posttraumatic stress, anxiety, and depression. This shows that as the magnitude of awareness about the infected individuals in the social vicinity increases, the perceived threat posed by the outbreak may increase and lead to significant psychological distress.

Most respondents (78.5%) reported that they spent over 2 hours per day watching TV and/or on social media. Increased daily exposure time to the media was significantly related to higher severity of posttraumatic stress, anxious and depressive symptoms in our sample. After controlling for the influence of sociodemographic and other variables, daily exposure time to the media predicted the increased PTSD symptoms (Table 4), but not the anxious or depressive symptoms (data not presented). The type of media being used was related to the anxiety severity in the preliminary analysis, however, this effect disappeared after controlling for the sociodemographic variables. Ultimately, prolonged exposure time to the distressing media content that covers sensitive material, and to the misinformation spreading through the social media networks might severely impact the psychological well-being of vulnerable individuals. This finding is consistent with recent studies (Yao, 2020; Chao et al., 2020).

During the lockdown period, job losses have been a continuously increasing problem around the world, with potentially serious psychosocial consequences. In our sample, a sizeable proportion of employees reported that their employment status changed after the COVID-19 outbreak. Overall, 18% of the employees reported that they had lost their job or were on temporary leave. Furthermore, 43% of the employees reported that they started working from home or paid infrequent office visits lately, and 7% reported that they were on paid leave. Only 30% of the employees did not report a significant change in their work routine. In multiple regression analysis, job loss or being on temporary leave during the COVID-19 outbreak was a positive predictor of increased PTSD and depressive symptoms.

As for the use of psychiatric services, our findings identified several levels of difficulties met by the patients throughout the outbreak. Even though the difficulty of getting an appointment at the outpatient clinic was low (6.6%), a significant proportion of patients (46.3%) reported that they had been avoiding the outpatient visits because of the fear of contracting the infection. Interestingly, avoiding the outpatient visits because of the fear of infection was significantly related to high psychological distress. Medication discontinuation (13.7%) and not being able to get the prescriptions filled (6.4%) during the outbreak were the other concerning findings. These indicate that difficulties in using the psychiatric services that arise from the fears of infection might be frequent, despite the seeming absence of serious problems in the regular services provided. Alternative routes such as telemedicine had been advocated to be potentially helpful in periods of social hardships, including the epidemics (Anthony, 2020; Fagherazzi et al., 2020; Yelloweess et al., 2020).

This study has a number of advantages. Our sample size was large, and the study was conducted around the peak of the outbreak, in one of the most heavily hit cities by the COVID-19 pandemic. However, there are several limitations to our study. First, we used a convenience sample, and caution must be exercised in generalization of our findings to the broader population of patients with psychiatric disorders. Second, this was a cross-sectional study which limits our ability to infer causality and also to conclude about the long term mental health consequences of the COVID-19 pandemic. Third, this study depended on self-report of patients instead of a structured clinical interview which could provide a better picture of the psychological distress in our patients. Finally, subjects in this study were mostly from an urban area, and this can limit generalization of the findings to subjects from rural environments.

In summary, patients with psychiatric disorders reported a high rate of psychological distress as an immediate response to the COVID-19 outbreak. In parallel, a significant number of patients described that their psychological status worsened during the outbreak. PTSD symptoms and comorbid depression, as well as anxiety, and impaired sleep comprise a substantial part of the distress described by these individuals. Various personal (i.e., age, educational attainment, gender, psychiatric diagnosis, chronic medical disease), behavioral (i.e., duration of media exposure) and social factors (i.e., infection prevalence in the social vicinity, and job loss during the outbreak) are likely to mediate the mental health effects in the context of COVID-19.

## Data Availability

The datasets generated during and/or analyzed during the current study are available from the corresponding on reasonable request.

## Acknowledgment

We thank our patients for their readiness to engage in this study.

## Contributors

Burç Çağrı Poyraz, Cana Aksoy Poyraz, Şenol Turan and Ömer Faruk Demirel designed the study and drafted the primary manuscript. Yasin Kavla, Ersel Bulu, İrem Hacısalihoğlu, Elif Burcu Ersungur supervised the recruitment and data management. Burç Çağrı Poyraz made the statistical analyses. All the authors had read and approved the final manuscript.

## References

Aydemir O., Guvenir T., Kuey L., Kultur S., 1997. The validity and reliability of the Turkish version of the Hospital Anxiety and Depression Scale Reliability. Turkish Journal of Psychiatry 8, 280–287.

Attepe Özden S., Tuncay T., 2018. The experiences of Turkish families caring for individuals with Schizophrenia: A qualitative inquiry. Int J Soc Psychiatry. 64(5),497–505. http://doi:10.1177/0020764018779090

Bjelland I., Dahl A.A., Haug T.T., Neckelmann D., 2002. The validity of the Hospital Anxiety and Depression Scale. An updated literature review. J Psychosom Res. 52(2), 69–77. http://doi:10.1016/s0022-3999(01)00296-3

Bocéréan C., Dupret E., 2014. A validation study of the Hospital Anxiety and Depression Scale (HADS) in a large sample of French employees. BMC Psychiatry 14,354. http://doi:10.1186/s12888-014-0354-0

Bokolo Anthony Jnr., 2020. Use of Telemedicine and Virtual Care for Remote Treatment in Response to COVID-19 Pandemic. J Med Syst. 44(7):132. http://doi:10.1007/s10916-020-015965

Brooks S.K., Webster R.K., Smith L.E., Woodland L., Wessely S., Greenberg N., Rubin G.J., 2020. The psychological impact of quarantine and how to reduce it: rapid review of the evidence. Lancet 395(10227), 912–920. http://doi:10.1016/S0140-6736(20)30460-8

Chao M., Xue D., Liu T., Yang H., Hall B.J., 2020. Media use and acute psychological outcomes during COVID-19 outbreak in China. J Anxiety Disord. 74:102248. http://doi:10.1016/j.janxdis.2020.102248

Creamer M., Bell R., Failla S., 2003. Psychometric properties of the Impact of Event Scale-Revised. Behav. Res. and Ther 41,1489–1496

Çorapçıoğlu A, Yargıç İ, Geyran P., Kocabaşoğlu N., 2006. Validity and Reliability of Turkish Version of “Impact of Event Scale-Revised” (IES-R). New Symposium Journal 44,14–22.

Dallel, S., Cancel, A., Fakra, E., 2018. Prevalence of Posttraumatic Stress Disorder in Schizophrenia Spectrum Disorders: A Systematic Review. Neuropsychiatry (London), 8(3), 1027–1037.

Djukanovic I., Carlsson J., Årestedt K., 2017. Is the Hospital Anxiety and Depression Scale (HADS) a valid measure in a general population 65-80 years old? A psychometric evaluation study. Health Qual Life Out. 15(1),193. http://doi:10.1186/s12955-017-0759-9

Fagherazzi G., Goetzinger C., Rashid M.A., Aguayo G.A., Huiart L., 2020. Digital Health Strategies to Fight COVID-19 Worldwide: Challenges, Recommendations, and a Call for Papers. J Med Internet Res. 22(6):e19284. http://doi:10.2196/19284

Hao F., Tan W., Jiang L., Zhang L., Zhao X., Zou Y., Hu Y., Luo X., Jiang X., McIntyre R.S., Tran B., Sun J.,Zhang Z., Ho R., Ho C., Tam W., 2020. Do psychiatric patients experience more psychiatric symptoms during COVID-19 pandemic and lockdown? A case-control study with service and research implications for immunopsychiatry. Brain Behav. Immun. 87,100–106. http:// doi:10.1016/j.bbi.2020.04.06

Iasevoli F., Fornaro M., D’Urso G., Galletta D., Casella C., Paternoster M., Buccelli C., de Bartolomeis A., the COVID-19 in Psychiatry Study Group., 2020. Psychological distress in patients with serious mental illness during the COVID-19 outbreak and one-month mass quarantine in Italy. Psychol Med,1-3. http://doi:10.1017/S0033291720001841

Lee, S.M., Kang, W.S., Cho, A.R., Kim, T., Park, J.K., 2018. Psychological impact of the 2015 MERS outbreak on hospital workers and quarantined hemodialysis patients. Compr. Psychiatry 87, 123–127. https://doi.org/10.1016/j.comppsych.2018.10.003.

Lommen M.J., Restifo K., 2009. Trauma and posttraumatic stress disorder (PTSD) in patients with schizophrenia or schizoaffective disorder. Community Ment Health J. 45(6):485–496. http://doi:10.1007/s10597-009-9248-x

Morina, N., Ehring, T., Priebe, S., 2013. Diagnostic Utility of the Impact of Event Scale– Revised in Two Samples of Survivors of War. PLoS ONE 8(12), e83916. Republic of Turkey Ministry of Health. (2020). https://covid19.saglik.gov.tr/

Sareen J., 2014. Posttraumatic stress disorder in adults: impact, comorbidity, risk factors, and treatment. Can J Psychiatry 59(9),460–467. http://doi:10.1177/070674371405900902

Sharma, V.K., Ho, C., 2020. A longitudinal study on the mental health of general population during the COVID-19 Epidemic in China. Brain Behav. Immun 87, 40–48. https://doi.org/10.1016/j.bbi.2020.04.028.

Strauss G.P., Duke L.A., Ross S.A., Allen D.N., 2011. Posttraumatic stress disorder and negative symptoms of schizophrenia. Schizophr Bull. 37(3), 603–610. http://doi:10.1093/schbul/sbp122

Tang B., Deng Q., Glik D., Dong J., Zhang L., 2017. A Meta-Analysis of Risk Factors for Post-Traumatic Stress Disorder (PTSD) in Adults and Children after Earthquakes. Int J Environ Res Public Health. 14(12);1537. http://doi:10.3390/ijerph14121537

Varshney M., Parel J.T., Raizada N., Sarin S.K., 2020. Initial psychological impact of COVID-19 and its correlates in Indian Community: An online (FEEL-COVID) survey. PLoS One 15(5), e0233874. https://doi:10.1371/journal.pone.0233874

Vindegaard N., Eriksen Benros M., 2020. COVID-19 pandemic and mental health consequences: Systematic review of the current evidence. Brain Behav. Immun. S0889-1591(20)30954–5. http://doi:10.1016/j.bbi.2020.05.048

Wang, C., Pan, R., Wan, X., Tan, Y., Xu, L., McIntyre, R.S., Choo, F., Tran, B., Ho, R., Sharma, V.K., Ho, C., 2020b. A longitudinal study on the mental health of general population during the COVID-19 Epidemic in China. Brain Behav. Immun. https://doi.org/10.1016/j.bbi.2020.04.028.

Weiss, D. S., Marmar, C. R., 1997. The Impact of Event Scale—Revised. In J. P. Wilson & T. M. Keane (Eds.), Assessing psychological trauma and PTSD (p. 399–411). Guilford Press Wikipedia contributors. (2020, June 20). COVID-19 pandemic in Turkey. In Wikipedia, The Free Encyclopedia. Retrieved 00:29, June 21, 2020, from https://en.wikipedia.org/wiki/COVID-19_pandemic_in_Turkey

Yao H., 2020. The more exposure to media information about COVID-19, the more distressed you will feel. Brain Behav. Immun 87,167–169. http://doi:10.1016/j.bbi.2020.05.031

Yelloweess P., Nakagawa K., Pakyurek M., Hanson A., Elder J., Kales H.C., 2020. Rapid Conversion of an Outpatient Psychiatric Clinic to a 100% Virtual Telepsychiatry Clinic in Response to COVID-19. Psychiatr Serv. http://doi:10.1176/appi.ps.202000230

Zhou J., Liu L., Xue P., Yang X., Tang X., 2020. Mental Health Response to the COVID-19 Outbreak in China. Am J Psychiatry 00:1-2.http://doi:10.1176/appi.ajp.2020.20030304

Zigmond A.S., Snaith R.P., 1983. The hospital anxiety and depression scale. Acta Psychiatr Scand 67(6),361–370. http://doi:10.1111/j.1600-0447.1983.tb09716.x

